# Ghost Gun Recovery and Firearm Deaths in California, 2014-2023

**DOI:** 10.1101/2025.09.08.25335327

**Authors:** Jemar R. Bather, Amanda I. Mauri, Zoe Lindenfeld, Saba Rouhani, Runhan Chen, Jinrui Fang, José A. Pagán, Melody S. Goodman, Diana Silver

**Author notes:** Corresponding author: Jemar R. Bather, Department of Biostatistics, NYU School of Global Public Health, 708 Broadway, New York, NY 10003, USA.

## Abstract

**Background:** Ghost guns are untraceable firearms assembled from online parts kits without background checks or waiting periods. Police nationally recovered 17 times more ghost guns in 2023 than 2017, yet the relationship between ghost gun recovery and firearm mortality is understudied. We investigated whether ghost gun recovery rates are significantly associated with subsequent firearm mortality rates across California’s 58 counties from 2014 to 2023.

**Methods:** We obtained yearly county-level data on ghost guns recovered in California from The Trace’s Gun Violence Data Hub, which aggregated data from the California Department of Justice’s October 2024 report *California’s Fight Against the Ghost Gun Crisis: Progress and New Challenges.* County-level firearm death counts (total, suicide, homicide) were pulled from the Centers for Disease Control and Prevention’s Restricted-Use Vital Statistics Data. Covariates included (1) urbanicity measured using the Rural-Urban Continuum Codes from the U.S. Department of Agriculture and (2) economic/racial segregation assessed by the Index of Concentration at the Extremes for income and race/ethnicity. We employed a hierarchical Bayesian approach to quantify the associations between ghost gun recoveries per capita and total firearm death rates in the following year. Exploratory analyses examined whether urbanicity and economic/racial segregation were significantly related to ghost gun recovery rates from 2014 to 2023.

**Results:** Controlling for urbanicity and economic/racial segregation, spatiotemporal models indicated that for every 20 ghost guns recovered per 100,000 population, there was an associated 5% increase in total firearm death rate (IRR: 1.05, 95% CrI: 1.02-1.08) and a 5% increase in firearm suicide rate (IRR: 1.05, 95% CrI: 1.02-1.09) in the following year. Ghost gun recovery rates were 298% higher in urban versus rural counties (IRR: 3.98, 95% CrI: 2.37-6.94) and increased 113% per 0.25-unit increase in economic/racial segregation score (IRR: 2.13, 95% CrI: 1.11-4.19).

**Conclusions:** This study provides the first empirical evidence examining the relationship between ghost gun recovery rates and subsequent increases in firearm mortality, particularly firearm suicides, across California counties from 2014 to 2023. Practitioners concentrating on suicide prevention efforts should be advised about the threat that ghost guns may present.

## Background

In 2022, over 48,000 people died due to firearms in the United States (U.S.), with 3,484 in California [1,2]. In California, 49% of these deaths were suicides, and 48% were homicides, and men were ten times more likely to die from firearm-related suicide than women [3].

Nationally, firearm mortality varies markedly across racial/ethnic lines, with rates among non-Hispanic Black (34.2 per 100,000) and American Indian/Alaska Native (20.7 per 100,000) subgroups higher than those of White (11.0 per 100,000), Native Hawaiian/Pacific Islander (10.1 per 100,000), Hispanic (8.7 per 100,000), and Asian (2.7 per 100,000) subgroups [3]. Firearm-related violence also imposes substantial economic consequences on the healthcare system [3,4]. In California, firearm-related violence is estimated to cost the health care system about $37 billion annually [3].

States vary substantially in their approach to firearm regulation, and rates of firearm violence vary dramatically across localities and states, with some research suggesting that stronger firearm restrictions may be associated with reductions in firearm deaths [5,6]. California has some of the strongest firearm laws in the country, and one of the lowest death rates from firearms [3,7]. Among the firearm regulations in California is a 2019 law that requires owners to obtain a unique serial number from the California Department of Justice for any “ghost gun” parts [8]. In addition, sales of firearm precursor parts, including unfinished receivers, must be conducted through licensed dealers [9]. However, enforcement remains difficult and largely relies on voluntary compliance. Research on the relationship between ghost gun availability and firearm deaths is limited [10–12]. Ghost guns may be particularly attractive to anyone prohibited from gun ownership, including persons with domestic violence histories or felony convictions, individuals with disqualifying mental health conditions, and minors [10,12]. Nationally, police recovered nearly 17 times more ghost guns in 2023 (n = 27,490) than in 2017 (n = 1,629), and the National Policing Project has highlighted the challenges for law enforcement that ghost guns pose [13,14]. While these numbers demonstrate a clear proliferation of ghost guns, research examining their public health impact remains limited and focused on a few cities, largely due to data availability.

Much of the literature on ghost guns has focused on California for two reasons [10,12]. First, California accounted for 55% of ghost guns recovered nationally from 2017 to 2021, according to the Bureau of Alcohol, Tobacco, Firearms, and Explosives [13]. Second, California is one of few states that generate extensive public law enforcement data [10,12]. Existing research studies have focused on only one or two cities, limiting generalizability to the broader state (where more resources may be available and where key policy decisions are also made) [10,12,15]. A study in Hayward, California found that about 5% of guns recovered in crimes were ghost guns 2015-2021, although in the years 2020 and 2021, the percentage was much higher [15]. Other studies found that ghost guns were more prevalent in violent and weapon-related offenses in Oakland and Los Angles but more prevalent in drug-related offenses in San Diego [10,12]. None of these studies examined the relationship between ghost guns and firearm-related mortality outcomes.

This study investigates the longitudinal relationship between ghost gun recovery rates and firearm deaths across 58 California counties from 2014 to 2023. We conceptualized ghost gun recovery rates as a proxy for illegal firearm availability and trafficking activity, both of which may increase firearm mortality [16]. We hypothesized that higher ghost gun recovery rates would be significantly associated with increased firearm deaths, measured as total firearm deaths and by specific types (e.g., firearm suicide, firearm homicide). We conducted an exploratory analysis to examine whether county-level socioeconomic factors were significantly associated with changes in ghost gun recovery patterns over time. We hypothesized that ghost gun recovery rates would be higher in urban areas and socially deprived neighborhoods, given the increased policing in these communities [17].

## Methods

### Data Source and Ghost Gun Recovery Measurement

We obtained yearly county-level data on ghost guns recovered in California from The Trace’s Gun Violence Data Hub [18], which aggregated data from the California Department of Justice’s October 2024 report *California’s Fight Against the Ghost Gun Crisis: Progress and New Challenges* [19] to create a dataset of ghost guns recovered in the state. The data comprised counts of guns used in crimes that were recovered by law enforcement and were determined by local police to be ghost guns across California’s 58 counties. For this study, ghost gun recovery rates per capita were defined as (number of ghost guns recovered/population size) × 100,000 using yearly county-level estimates from the American Community Survey. This study did not require review by the New York University Institutional Review Board and followed the Strengthening the Reporting of Observational Studies in Epidemiology guidelines [20].

### Firearm Mortality

We obtained county-level firearm death counts from the Centers for Disease Control and Prevention’s (CDC) Restricted-Use Vital Statistics Data [21]. Through this system, each state sends the CDC death certificates that contain the underlying cause of death using the International Classification of Diseases, 10^th^ Revision coding methods. The outcomes of interest were the total count of firearm-related deaths, suicides, and homicides from 2014 to 2023. Given that firearm-related suicide is more common among males than females [22], we also stratified the suicide outcome by sex. Unintentional firearm deaths and undetermined intent firearm deaths were included in the total count but not analyzed as separate outcomes as they comprise a very low percentage of annual total firearm-related deaths. Additional details on the death reporting protocols are available on the CDC’s website and in prior studies [21,23,24]. Access to the Restricted-Use Vital Statistics Data was approved by CDC’s National Center for Health Statistics.

### Covariates

Covariates included urbanicity and economic/racial segregation. Each county was classified as urban or rural using the Rural-Urban Continuum Codes from the U.S. Department of Agriculture [25]. Economic/racial segregation was measured using the Index of Concentration at the Extremes (ICE) for income and race/ethnicity [26,27]. This index quantifies the extent to which county residents are concentrated at racial and economic extremes, comparing areas with high deprivation (Black residents, including Hispanic, in the 20^th^ income percentile) to areas with high privilege (non-Hispanic White residents in the 80^th^ income percentile). Based on one-year American Community Survey estimates, ICE values range from –1 to 1, where –1 indicates the maximum concentration of deprived populations and 1 indicates the maximum concentration of privileged populations. We multiplied ICE values by –1 so that higher scores indicate a greater concentration of deprived populations. This panel dataset contained no missing values across all counties and variables.

### Statistical Analysis

For the primary analysis, we employed a hierarchical Bayesian approach to examine the relationship between temporal changes in ghost gun recoveries per capita and total firearm death rates in the following year. To establish temporal precedence, we lagged ghost gun recovery rates by one year, ensuring that the exposure (ghost gun recovery rates) preceded the outcome (total firearm death rates). This model controlled for county characteristics measured in 2013: urbanicity and economic/racial segregation.

This hierarchical framework was selected to control for spatial and temporal correlations in county-level data [28]. We used negative binomial regression to account for potential overdispersion in total firearm death counts. We incorporated several key components into the model, including an offset term (logarithm of the population size) to account for varying county populations; a spatial random effect based on the Besag-York-Mollié 2 model to capture structured and unstructured heterogeneity between neighboring counties [29]; and a temporal random effect using a first-order random walk to allow for non-linear trends over time [30]. We applied this same modeling framework in secondary analyses to examine associations between ghost gun recovery rates and specific types of firearm deaths: suicides overall and stratified by sex, and homicides. Additionally, we conducted exploratory analyses to investigate how county-level factors (urban/rural status [time-invariant] and economic/racial segregation [time-varying]) were associated with ghost gun recovery rates from 2014 to 2023.

All models were estimated using integrated nested Laplace approximation [31]. We tabulated incidence rate ratios (IRRs) and 95% credible intervals (CrIs), with CrIs not including one determining statistical significance. All analyses were performed using R version 4.5.1 (R Core Team, R Foundation for Statistical Computing).

## Results

### Descriptive Statistics

Table 1 compares all study variables by year in California’s 58 counties. From 2014 to 2023, average county-level firearm deaths increased: total firearm deaths grew 9.7% (from 34.78 to 38.14), firearm suicides rose 8.2% (from 19.93 to 21.57), and firearm homicides increased 10.5% (from 14.17 to 15.66). Suicide rates by sex exhibited differential patterns as male suicide rates grew 11.0% (from 16.81 to 18.66), and female suicide rates decreased by 6.7% (from 3.12 to 2.91).

**Table 1.**
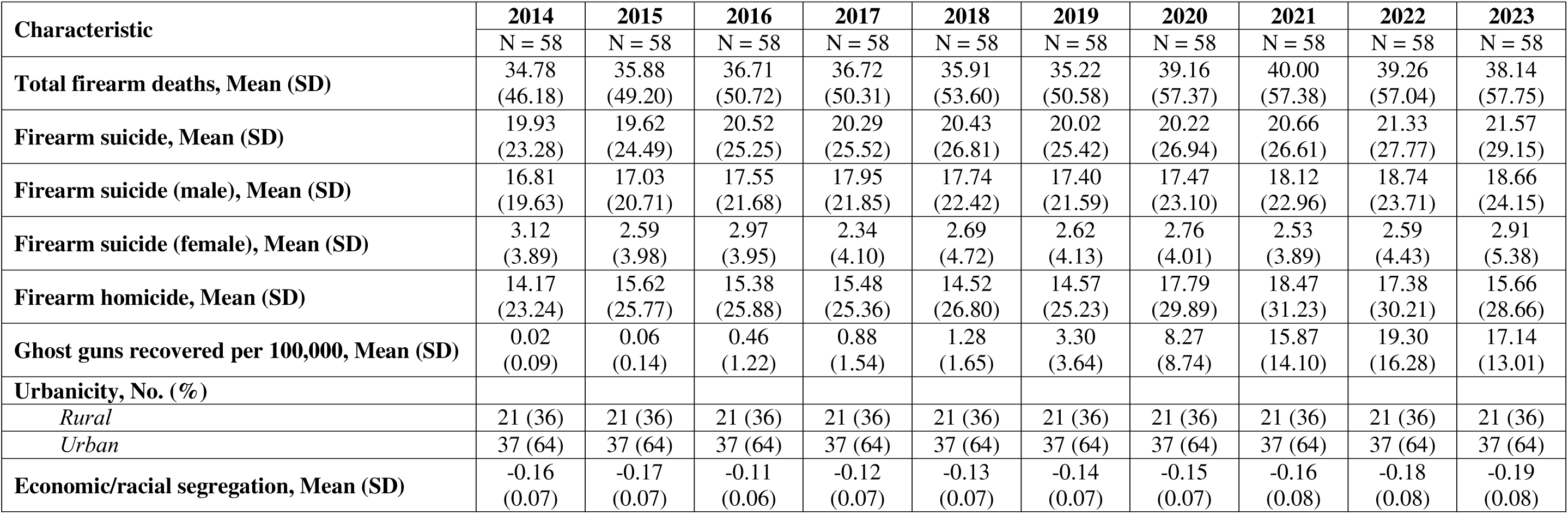
Trends in firearm mortality and related characteristics across California’s 58 counties, 2014-2023.

| **Characteristic** | **2014**<br>N = 58 | **2015**<br>N = 58 | **2016**<br>N = 58 | **2017**<br>N = 58 | **2018**<br>N = 58 | **2019**<br>N = 58 | **2020**<br>N = 58 | **2021**<br>N = 58 | **2022**<br>N = 58 | **2023**<br>N = 58 |
| --- | --- | --- | --- | --- | --- | --- | --- | --- | --- | --- |
| **Total firearm deaths, Mean (SD)** | 34.78<br>(46.18) | 35.88<br>(49.20) | 36.71<br>(50.72) | 36.72<br>(50.31) | 35.91<br>(53.60) | 35.22<br>(50.58) | 39.16<br>(57.37) | 40.00<br>(57.38) | 39.26<br>(57.04) | 38.14<br>(57.75) |
| **Firearm suicide, Mean (SD)** | 19.93<br>(23.28) | 19.62<br>(24.49) | 20.52<br>(25.25) | 20.29<br>(25.52) | 20.43<br>(26.81) | 20.02<br>(25.42) | 20.22<br>(26.94) | 20.66<br>(26.61) | 21.33<br>(27.77) | 21.57<br>(29.15) |
| **Firearm suicide (male), Mean (SD)** | 16.81<br>(19.63) | 17.03<br>(20.71) | 17.55<br>(21.68) | 17.95<br>(21.85) | 17.74<br>(22.42) | 17.40<br>(21.59) | 17.47<br>(23.10) | 18.12<br>(22.96) | 18.74<br>(23.71) | 18.66<br>(24.15) |
| **Firearm suicide (female), Mean (SD)** | 3.12<br>(3.89) | 2.59<br>(3.98) | 2.97<br>(3.95) | 2.34<br>(4.10) | 2.69<br>(4.72) | 2.62<br>(4.13) | 2.76<br>(4.01) | 2.53<br>(3.89) | 2.59<br>(4.43) | 2.91<br>(5.38) |
| **Firearm homicide, Mean (SD)** | 14.17<br>(23.24) | 15.62<br>(25.77) | 15.38<br>(25.88) | 15.48<br>(25.36) | 14.52<br>(26.80) | 14.57<br>(25.23) | 17.79<br>(29.89) | 18.47<br>(31.23) | 17.38<br>(30.21) | 15.66<br>(28.66) |
| **Ghost guns recovered per 100,000, Mean (SD)** | 0.02<br>(0.09) | 0.06<br>(0.14) | 0.46<br>(1.22) | 0.88<br>(1.54) | 1.28<br>(1.65) | 3.30<br>(3.64) | 8.27<br>(8.74) | 15.87<br>(14.10) | 19.30<br>(16.28) | 17.14<br>(13.01) |
| **Urbanicity, No. (%)** |  |  |  |  |  |  |  |  |  |  |
| *Rural* | 21 (36) | 21 (36) | 21 (36) | 21 (36) | 21 (36) | 21 (36) | 21 (36) | 21 (36) | 21 (36) | 21 (36) |
| *Urban* | 37 (64) | 37 (64) | 37 (64) | 37 (64) | 37 (64) | 37 (64) | 37 (64) | 37 (64) | 37 (64) | 37 (64) |
| **Economic/racial segregation, Mean (SD)** | -0.16<br>(0.07) | -0.17<br>(0.07) | -0.11<br>(0.06) | -0.12<br>(0.07) | -0.13<br>(0.07) | -0.14<br>(0.07) | -0.15<br>(0.07) | -0.16<br>(0.08) | -0.18<br>(0.08) | -0.19<br>(0.08) |

Table 2 presents the results from the primary and secondary analyses testing whether increases in ghost gun recovery rates were significantly related to rates of total and specific types of firearm deaths in the following year. Spatiotemporal models indicated that for every 20 ghost guns recovered per 100,000 population, there was an associated 5% increase in total firearm death rate (IRR: 1.05, 95% CrI: 1.02, 1.08), and a 5% increase in firearm suicide rate (IRR: 1.05, 95% CrI: 1.02, 1.09) in the following year. A similar pattern emerged for male firearm suicide rates (IRR: 1.05, 95% CrI: 1.01, 1.09) but not for female suicide rates (IRR: 1.02, 95% CrI: 0.93, 1.11). We found no evidence of a significant association between ghost gun recovery and firearm homicide rates (IRR: 1.02, 95% CrI: 0.95, 1.10). All models controlled for urbanicity and economic/racial segregation measured in 2013.

**Table 2.** Adjusted associations of ghost gun recovery rates with firearm death rates across California’s 58 counties, 2014-2023.

|  | **Total firearm deaths** | **Firearm suicide** |  |  | **Firearm homicide** |
| --- | --- | --- | --- | --- | --- |
|  |  | **Total** | **Male** | **Female** |  |
|  | IRR (95% CrI) | IRR (95% CrI) | IRR (95% CrI) | IRR (95% CrI) | IRR (95% CrI) |
| Ghost guns recovered per 100k (20-unit increase) | 1.05 (1.02, 1.08) | 1.05 (1.02, 1.09) | 1.05 (1.01, 1.09) | 1.02 (0.93, 1.11) | 1.02 (0.95, 1.10) |
| IRR = incidence rate ratio; CrI = credible interval |  |  |  |  |  |
| Ghost gun recovery rates were scaled to represent 20-unit increases per 100,000 population. |  |  |  |  |  |
| All models controlled for covariates measured in 2013: urbanicity & economic/racial segregation. |  |  |  |  |  |

Exploratory analyses examined factors associated with ghost gun recovery rates (Table 3). Over the 2014-2023 period, ghost gun recovery rates were almost 4-fold higher (IRR: 3.98, 95% CrI: 2.37-6.94) in urban counties compared to rural counties. Counties with higher economic/racial segregation (per 0.25-point increase in the ICE score) had 113% higher ghost gun recovery rates (IRR: 2.13, 95% CrI: 1.11-4.19) than counties with lower economic/racial segregation.

**Table 3.** Associations of urbanicity and economic/racial segregation with ghost gun recovery rates across California’s 58 counties, 2014-2023.

|  | **Incidence Rate**<br>**Ratio** | **Credible**<br>**Interval** |
| --- | --- | --- |
| **Urbanicity** |  |  |
| Rural (reference) |  |  |
| Urban | 3.98 | (2.37, 6.94) |
| **Economic/racial segregation** | 2.13 | (1.11, 4.19) |
| Economic/racial segregation scores were scaled to represent 0.25-unit increases. |  |  |
| Urbanicity was time-invariant and economic/racial segregation was time-varying. |  |  |

## Discussion

Ghost guns are emerging as a critical public health threat in the U.S., yet little is known about their relationship with firearm mortality [10–12]. Drawing on data from a newly available source, we quantified the association between ghost gun recovery rates and firearm deaths across 58 California counties from 2014 to 2023. Results from the spatiotemporal analysis demonstrated that for every 20 additional ghost guns recovered per 100,000 population, there was a 5% relative increase in the total firearm death rate in the following year. This association was also significant for firearm suicide rates but not for firearm homicides. Additionally, exploratory analyses indicated that counties with urban classification and higher levels of economic/racial segregation experienced significantly higher ghost gun recovery rates.

Collectively, these findings advance ghost gun research and policy discussions while providing data-driven insights for law enforcement and firearm violence prevention organizations.

The observed associations between ghost gun recoveries and firearm mortality are likely conservative estimates. The data captured only ghost guns retrieved by law enforcement and not those remaining in circulation, thereby potentially underestimating the true ghost gun prevalence. This measurement error may have biased our effect estimates toward the null [32]. Therefore, the actual relationship between ghost gun availability and firearm mortality may be stronger than our results suggest. Even with this potential underestimation, we found that ghost gun recoveries were significantly associated with total firearm death rates in the following year. This association was more pronounced for firearm suicides than for homicides. Our finding that the relationship persisted when suicide was stratified by sex confirms other studies that have noted the relationship between firearms and suicides, particularly among men [33,34]. Future research should determine whether those who are unable to obtain a gun due to felon status, mental illness, or substance use disorders turn to ghost guns. Wintemute called for a number of actions to regulate ghost guns, including calling for state and federal governments to prohibit the manufacture of ghost guns, to forbid persons who violate laws pertaining to ghost guns from owning any gun, and for greater clarity from the Bureau of Alcohol, Tobacco, Firearms, and Explosives regarding firearm precursors [11]. Our study adds to the imperative to address these policy ideas [11].

The exploratory analyses indicated that ghost gun recovery rates varied considerably by rural-urban status and levels of economic/racial segregation. This finding aligns with extensive evidence on differential policing in deprived neighborhoods [35–40]. Studies have demonstrated that individuals in these communities face higher rates of police stops [38,40], searches [38], arrests [37], and fatal police encounters [35,36,39] compared to their counterparts in privileged neighborhoods. Scholars have conceptualized this pattern through the minority threat hypothesis, which posits that police perceive deprived neighborhoods as requiring more intensive enforcement [41,42]. Given these policing patterns, it is plausible that the higher ghost gun recovery rates in urban areas and communities with high economic/racial deprivation reflect more frequent police contact and searches in these communities, as opposed to greater illegal firearm availability and trafficking activity. Accordingly, future work should investigate whether geographic variations in recovery rates reflect true differences in ghost gun prevalence or differential enforcement patterns.

The current study had several notable strengths. First, the restricted-access mortality data provided granular outcome measurement, avoiding the left-censoring that occurs when areas have fewer than ten deaths [43]. Second, we leveraged a novel data source capturing ghost gun recoveries across all 58 California counties over ten years (2014-2023), extending beyond the city-specific analyses in prior research [10,12,15]. Third, the spatiotemporal modeling approach addressed two key sources of correlation. The spatial random effect accounted for similarities between neighboring counties, and the temporal random effect controlled for statewide trends that affected all counties. This comprehensive analytic framework allowed us to better isolate the association between ghost gun recoveries and firearm mortality while accounting for geographic clustering and temporal patterns.

Despite these strengths, several limitations warrant consideration. With a focus on California, this study’s findings may not generalize to states with different firearm regulations and enforcement practices. As such, future research should replicate these analyses in other U.S. states and territories [44], as well as nationally. Although we constructed a longitudinal design, unmeasured time-varying confounders may have influenced the observed relationships. Changes in police funding, violence intervention programs, or community-level factors could affect ghost gun recoveries and firearm mortality [45–48]. Another consideration is that the ecological design precluded individual-level inferences about ghost gun possession and mortality risk [32]. Finally, while we demonstrated that areas with higher ghost gun recovery rates experienced increased rates of total firearm deaths and firearm suicides, we could not determine whether the recovered ghost guns were the actual weapons used in these deaths.

## Conclusions

This study provides the first empirical evidence examining the relationship between ghost gun recovery rates and subsequent increases in overall firearm mortality and firearm suicides, across California counties from 2014 to 2023. Practitioners working with those at high risk of mental illness should be aware that the availability of ghost guns may subvert regular channels for gun oversight. Policymakers concerned about addressing suicide could address the need for greater regulation of ghost guns, by working with federal, state, and local authorities. Law enforcement may want to increase enforcement of ghost gun regulations, especially in rural areas where suicide rates are often highest [49,50]. Last, ghost gun recovery rates were significantly higher in urban areas and communities experiencing greater economic/racial deprivation, illustrating potential geographic and socioeconomic differences in exposure to these untraceable weapons.

## Declarations

### Ethics approval and consent to participate

This study did not require IRB approval because the New York University Institutional Review Board did not consider it human subjects research. Therefore, written informed consent was not required. This study was performed in accordance with the ethical standards laid down in the 1964 Declaration of Helsinki and its later amendments or comparable ethical standards.

## Consent for publication

Not applicable.

## Availability of data and material

The ghost gun data are openly available (https://datahub.thetrace.org/dataset/california-ghost-guns-stolen-guns-and-more/).

## Authors contributions

**Jemar R. Bather:** Conceptualization, Methodology, Software, Formal analysis, Investigation, Data Curation, Writing – Original Draft, Project administration. **Amanda I. Mauri:** Investigation, Data Curation, Writing – Review & Editing. **Zoe Lindenfeld:** Investigation, Data Curation, Writing – Review & Editing. **Saba Rouhani:** Investigation, Writing – Review & Editing. **Runhan Chen:** Validation, Investigation, Data Curation, Writing – Review & Editing. **Jinrui Fang:** Validation, Investigation, Data Curation, Writing – Review & Editing. **José A. Pagán:** Investigation, Writing – Review & Editing. **Melody S. Goodman:** Investigation, Resources, Writing – Review & Editing. **Diana Silver:** Validation, Investigation, Writing – Review & Editing.

## Abbreviations

CDC: Centers for Disease Control and Prevention
ICE: Index of Concentration at the Extremes
IRR: Incidence Rate Ratio
CrI: Credible Interval

## Acknowledgements

We thank Samantha Storey, George LeVines, Ava Sasani, Aaron Mendelson, the Gun Violence Data Hub, and The Trace for making the ghost guns database publicly available.

